# A Neuropathology Case Report of a Woman with Down Syndrome who Remained Cognitively Stable

**DOI:** 10.1101/2024.06.01.24308050

**Authors:** Jr-Jiun Liou, Jerry Lou, Jamie Nakagiri, William Yong, Christy L. Hom, Eric W. Doran, Minodora Totoiu, Ira Lott, Mark Mapstone, David B. Keator, Adam M. Brickman, Sierra Wright, Brittany Nelson, Florence Lai, Laura Xicota, Lam-Ha T. Dang, Jinghang Li, Tales Santini, Joseph M. Mettenburg, Milos D. Ikonomovic, Julia Kofler, Tamer Ibrahim, Elizabeth Head, the Alzheimer Biomarker Consortium - Down Syndrome

## Abstract

In this neuropathology case report, we present findings from an individual with Down syndrome (DS) who remained cognitively stable despite Alzheimer’s disease (AD) neuropathology. Clinical assessments, fluid biomarkers, neuroimaging, and neuropathological examinations were conducted to characterize her condition. Notably, her ApoE genotype was E2/3, which is associated with a decreased risk of dementia. Neuroimaging revealed stable yet elevated amyloid profiles and moderately elevated tau levels, while neuropathology indicated intermediate AD neuropathologic change with Lewy body pathology and cerebrovascular pathology. Despite the presence of AD pathology, the participant demonstrated intact cognitive functioning, potentially attributed to factors such as genetic variations, cognitive resilience, and environmental enrichment. The findings suggest a dissociation between clinical symptoms and neuropathological changes, emphasizing the complexity of AD progression in DS. Further investigation into factors influencing cognitive resilience in individuals with DS, including comorbidities and social functioning, is warranted. Understanding the mechanisms underlying cognitive stability in DS could offer insights into resilience to AD neuropathology in people with DS and in the general population and inform future interventions.

## Introduction

Aging adults with Down syndrome (DS) accumulate Alzheimer’s disease (AD) neuropathology, including beta-amyloid plaques and neurofibrillary tangles, by the age of 40 years [1, 2]. Most older people with DS also develop mild cognitive impairment (MCI) or early signs of dementia between 48 and 56 years of age [3-10]. The Alzheimer Biomarker Consortium-Down Syndrome (ABC-DS) was established in 2015 with the goal to characterize biomarkers of AD in people with DS [11]. In this case report we describe a participant in ABC-DS who remained cognitively stable at an age where up to 90% of people with DS develop the clinical features of MCI or dementia [12-14].

## Methods

### Case description

We describe a woman in her 60’s with ApoE2/3 and trisomy 21 who remained cognitively stable up until her death. The participant was followed for 9 years in two NIH funded longitudinal studies of AD in DS before enrolling in the ABC-DS. Institutional Review Board approval and informed consent from the participant were obtained.

### Clinical assessment

Full scale IQ (FSIQ) was acquired from the medical records, where assessments were conducted by a community provider using the Wechsler Adult Intelligence Scale, Revised Edition (WAIS-R) [15] at two time points in her 30’s. The Kaufman Brief Intelligence Test, 2^nd^ Edition (KBIT-2; [16]) was administered to the participant when she was in her 60’s by an ABC-DS clinician. Overall cognitive function was assessed with the Down Syndrome Mental Status Examination (DSMSE) [17] and the Rapid Assessment for Developmental Disabilities, 2^nd^ Edition [18]. Memory was evaluated with the Modified Cued Recall test [19]. Attention and aspects of executive function were assessed with the Cats and Dogs Stroop [20] Naming and Switch tasks. Additionally, language, visuospatial abilities, and motor performance were evaluated. Dementia symptoms were assessed and characterized as the sum of cognitive and social scores from the Dementia Questionnaire for People with Learning Disabilities (DLD) [21]. The consensus diagnosis, based only on clinical and neuropsychological data, at both time points when the participant was in her 60’s was cognitively stable [11].

### Fluid biomarkers

Cerebrospinal fluid (CSF) was collected from the participant following protocols consistent with previous studies [22]. The participant underwent a lumbar puncture procedure, from which approximately 10-20 mL of CSF was collected through gravity drip. Subsequently, the CSF samples were flash-frozen on dry ice before being shipped to the Fluid Biomarker Core at Washington University in St. Louis. Upon receipt, the samples were thawed and aliquoted into polypropylene tubes before being stored at –80°C. CSF biomarkers, including amyloid beta 40 (Aβ40), amyloid beta 42 (Aβ42), total tau (tTau), and phospho-tau181 (pTau181), were measured using an automated immunoassay LUMIPULSEG1200 (Fujirebio, USA).

Peripheral blood samples were collected from the participant, with protocols for processing and analysis similar to previous studies [23]. Plasma biomarkers, including Aβ40, Aβ42, tTau, pTau181, phospho-tau217 (pTau217), glial fibrillary acidic protein (GFAP), and neurofilament light chain (Nf-L), were measured. Plasma Aβ40, Aβ42, GFAP, and Nf-L as well as pTau181 were analyzed using Simoa Neurology 4-Plex E and Simoa p-Tau 181 assays (Quanterix, USA), respectively [24]. Plasma p-tau217 concentration was measured using an immunoassay on a Mesoscale Discovery platform developed by Lilly Research Laboratories, calibrated with a synthetic p-tau217 peptide [25, 26]. All fluid biomarker analyses were conducted by staff blinded to the clinical and imaging data.

### Genotyping

Genomic DNA was extracted from peripheral blood collected and genotyped using Infinum General Screening Array V2, as previously described [27]. Mosaicism was assessed plotting B allele frequencies against position on chromosome 21 using GenomeStudio (Illumina, San Diego, CA, USA). Deviations of the expected 0, 0.3, 0.6, 1 B allele frequencies, were considered to show mosaicism [28].

### Neuroimaging

Four ^18^F-AV-45 (florbetapir; Amyloid) and two ^18^F-AV-1451 (flortaucipir; Tau) scans were conducted during the last 5 years of the participant’s life using the High Resolution Research Tomograph (HRRT; Siemens; orientation=axial, voxel size=1.2mm^3^, matrix size=256×256×207, reconstruction=OP-OSEM3D). The participant was injected with 10 ± 1.0 mCi of florbetapir or flortaucipir with an uptake time of ∼40 minutes for florbetapir and ∼70 minutes for flortaucipir while at rest. Image acquisition followed the Alzheimer’s Disease Neuroimaging Initiative (ADNI) [29] protocol consisting of 4x5 minute frames collected 50-70 minutes (florbetapir) or 80-100 minutes (flortaucipir) after injection of the ligand. The PET frames were realigned, averaged, and co-registered with their respective MRI scans. MRI segmentations were computed with FreeSurfer (FS6; RRID:SCR_001847). Regions of interest (ROI) were extracted in the native MRI space from the FS6 Desikan/Killiany atlas [30] segmentations. The PET counts were converted to standardized uptake value ratio (SUVR) units using the cerebellum cortex reference region. Correction for partial volume effects was performed using PETSurfer [31]. ROI average values were calculated for meta-regions corresponding to each Braak staging (I-VI) region using the temporally closest MR scan FreeSurfer segmentations [32]. Antemortem MR scans at 3T were conducted using either Philips Achieva or Siemens Prisma scanner with a body coil . The scan protocol included T1-weighted MPRAGE at 1.0 mm resolution, T2-weighted FLAIR at 5.0 mm resolution, and T2-star GRE at 4.0 mm resolution.

### Postmortem MRI

The use of autopsy tissue for research was approved by the Committee for Oversight of Research and Clinical Training Involving Decedents at the University of Pittsburgh and the University of California, Irvine. The postmortem interval was 15 hours. The left hemisphere was fixed in 4% paraformaldehyde for three weeks and subsequently embedded in 1.5% (w/v) agar (Millipore Sigma, A5431) and 30% sucrose (Fisher, S5-3) hydrogel [33] in a 3D printed container [34-36] for postmortem MRI scanning. This scanning process utilized a 7T human MRI scanner (Siemens, Germany), featuring a custom-built radiofrequency Tic-Tac-Toe head coil system equipped with 16 transmit channels and 32 receive channels [37-39]. Structural imaging encompassed the acquisition of T1-weighted MP2RAGE scans at a resolution of 0.37 mm isotropic, T2-weighted SPACE imaging at a resolution of 0.41 mm, and T2-star GRE (susceptibility-weighted imaging, SWI) at a resolution of 0.37 mm, specifically for detecting cerebral microhemorrhages. Postmortem T1 scans were employed to align antemortem T1 and gross images, facilitating comprehensive analysis.

### Neuropathology

The right hemisphere was frozen, while the left hemisphere was fixed and embedded for postmortem MRI and subsequent histology. Tissue sampling and staining included all brain regions of the left hemisphere recommended by the 2012 National Institutes of Aging – Alzheimer’s Association consensus criteria for the neuropathological evaluation of Alzheimer’s disease [40, 41].

Immunohistochemical staining for beta-amyloid (Biolegend #803015, 1:1000) was performed to characterize the Thal stage [42]. Tau staining (Agilent #A0024, 1:3000) was performed to determine Braak stage [43] to assess neuritic plaque density by Consortium to Establish a Registry for Alzheimer’s disease (CERAD) criteria [44]. According to the fourth consensus report of the dementia with Lewy bodies consortium [45], alpha-synuclein staining (Millipore Sigma #AB5038, 1:1000) was used to detect the presence of Lewy body-related pathology. A non-phospho-TDP-43 antibody (Proteintech #10782-2-AP, 1:2000) was used to detect TDP-43 signal in the amygdala, hippocampus and/or entorhinal/inferior temporal cortex, as well as neocortex.

## Results

### Clinical assessment

The participant had an IQ of 69 indicating a mild level of intellectual disability (Table 1). Her body mass index (BMI) was recorded at 36.1, indicating obesity. Although her gait was slow with short steps, it was likely due to her osteoarthritis of bilateral knees and joint pain.

**Table 1.** Clinical data of the case.

|  |  | Visit 1 | Visit 2 | Visit 3 | Visit 4 | Visit 5 | Visit 6 | Visit 7 | Visit 8 |
| --- | --- | --- | --- | --- | --- | --- | --- | --- | --- |
| Cognition | FSIQ | WAIS-R=69 | - | - | - | - | - | - | KBIT-2=64 |
|  | DSMSE | - | 84 | 88 | 90.5 | 84 | 85.5 | 91 | 83 |
| Memory | Cued Recall | - | - | - | - | 36 | 36 | 35 | 30 |
| Executive function | Cats and Dogs Stroop Naming Time | - | - | - | - | 11.5" | 9.6" | 10.3" | 11.7" |
|  | Cats and Dogs Stroop Naming Errors | - | - | - | - | 0 | 0 | 0 | 0 |
|  | Cats and Dogs Stroop Switch Time | - | - | - | - | 21.1" | 20.1" | 24.6" | 20.3" |
|  | Cats and Dogs Stroop Switch Errors | - | - | - | - | 0 | 0 | 1 | 0 |
| Visuospatial | Block Design | - | 19 | 15 | 21 | 22 | 18 | 20 | 18 |
|  | VMI | - | 22 | 22 | 22 | 23 | 22 | 22 | 19 |
| Motor | Tinetti Gait | - | - | - | - | 12 | 12 | 11 | 10 |
| Dementia symptoms | DLD Cognitive | - | 1 | 0 | 0 | 0 | 0 | 7 | 0 |
|  | DLD Social | - | 4 | 4 | 1 | 6 | 10 | 22 | 0 |

The participant received private school education from childhood through adolescence at an institute licensed to educate students with intellectual disabilities and behavioral challenges, something that the majority of children with DS from her generation would not have received. This enriched learning environment may have contributed to her cognitive reserve.

Until her passing, she maintained cognitive stability across visits, as evidenced by consistent performance on cognitive tests such as the DSMSE. Her Cued Recall memory scores remained stable, and she demonstrated minimal impairment in executive function, indicated by consistent Cats and Dogs Stroop Naming scores with no errors over recent years before her passing. Notably, her DLD scores in both cognitive and social domains were remarkably low, registering zero. Furthermore, she retained functional capacity, managing most of her own cooking and shopping until her death. Most of the clinical team arrived at a consensus diagnosis of cognitively stable at every cycle, although there was worsening in the DLD social scores showing increases, reflecting onset of mild behavioral or psychiatric symptoms. However, she was reported to have appropriate social engagement and behaviors at the last visit prior to her death.

### Fluid biomarkers

Analysis of CSF protein concentrations (Table 2) revealed elevated concentrations of Aβ40, resulting in a lower Aβ42/40 ratio. Levels of CSF pTau181 and tTau exceeded the manufacturer cutoffs established in the Amsterdam Dementia Cohort, which includes individuals with subjective cognitive decline, MCI, AD, and other forms of dementia [46-48]. However, it is worth noting that these levels were lower than the average observed in our previous study involving 341 participants with DS, similarly aged carriers of autosomal dominant AD mutations, and non-carrier siblings [22]. Furthermore, her plasma levels of tTau and GFAP exceeded those reported in published studies [23, 49, 50].

**Table 2.**
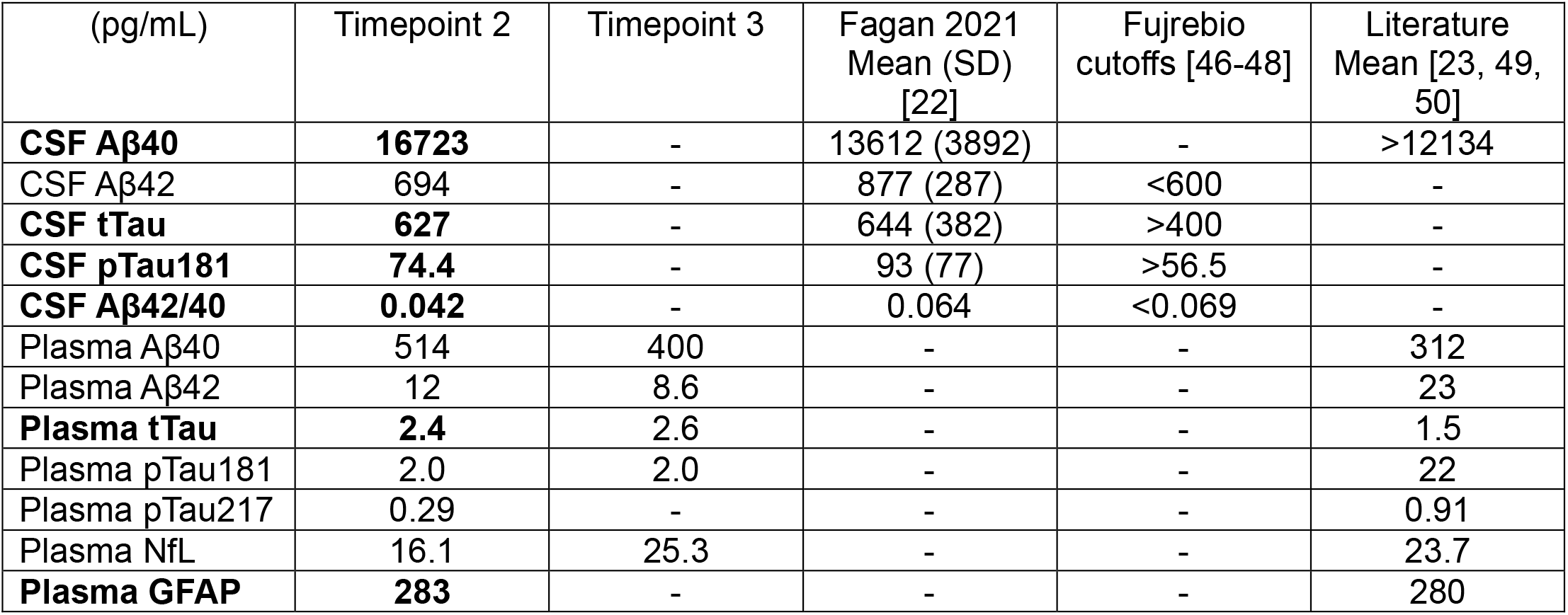
CSF and plasma biomarkers of the case. **Numbers above or below the average are bolded**.

| (pg/mL) | Timepoint 2 | Timepoint 3 | Fagan 2021<br>Mean (SD)<br>[22] | Fujrebio<br>cutoffs [46-48] | Literature<br>Mean [23, 49,<br>50] |
| --- | --- | --- | --- | --- | --- |
| **CSF Aβ40** | **16723** | - | 13612 (3892) | - | >12134 |
| CSF Aβ42 | 694 | - | 877 (287) | <600 | - |
| **CSF tTau** | **627** | - | 644 (382) | >400 | - |
| **CSF pTau181** | **74.4** | - | 93 (77) | >56.5 | - |
| **CSF Aβ42/40** | **0.042** | - | 0.064 | <0.069 | - |
| Plasma Aβ40 | 514 | 400 | - | - | 312 |
| Plasma Aβ42 | 12 | 8.6 | - | - | 23 |
| **Plasma tTau** | **2.4** | 2.6 | - | - | 1.5 |
| Plasma pTau181 | 2.0 | 2.0 | - | - | 22 |
| Plasma pTau217 | 0.29 | - | - | - | 0.91 |
| Plasma NfL | 16.1 | 25.3 | - | - | 23.7 |
| **Plasma GFAP** | **283** | - | - | - | 280 |

### Genotyping

The B allele distribution obtained from the genome-wide association study (GWAS) data suggests the presence of approximately 10% disomic cells. This indicates mosaicism at the time of peripheral blood collection.

### Neuroimaging

In evaluating the Amyloid PET ROI averages in the PET amyloid-specific regions, we found stable, yet elevated (SUVR range 1.0-1.7), amyloid profiles at which time the SUVR values increased with PET amyloid-specific stage IV regions (inferior temporal cortex; middle temporal cortex; temporal pole; thalamus; caudal, rostral, isthmus, posterior cingulate; insula) yielding the largest increases between timepoints 3-4 and PET amyloid-specific stage V regions (frontal cortex; parietal cortex; occipital cortex; transverse, superior temporal cortex; precuneus; banks of superior temporal sulcus; nucleus accumbens caudate nucleus; putnam) yielding the highest SUVR of 1.8. These increases can be seen in Figure 1A, showing elevated Amyloid. Further, we see the same profile in superior frontal, rostral middle frontal, inferior parietal, and posterior cingulate regions. The magnitude of Amyloid SUVR load in her final scan in the regions evaluated appears to be consistent with levels seen in DS with MCI as described in Keator et al. [51]. In contrast, tau PET ROIs of PET tau-specific regions were flat across the two timepoints with SUVRs in the range of 1.0 – 1.4, indicating moderately elevated tau, yet stable. Antemortem MRI T1-weighted MPRAGE indicated minimal brain atrophy and numerous unusual spherical lesions primarily involving the white matter and to a lesser extent the region of the caudate nuclei; the central portion of the lesions follow CSF signal on all sequences.

**Figure 1.**
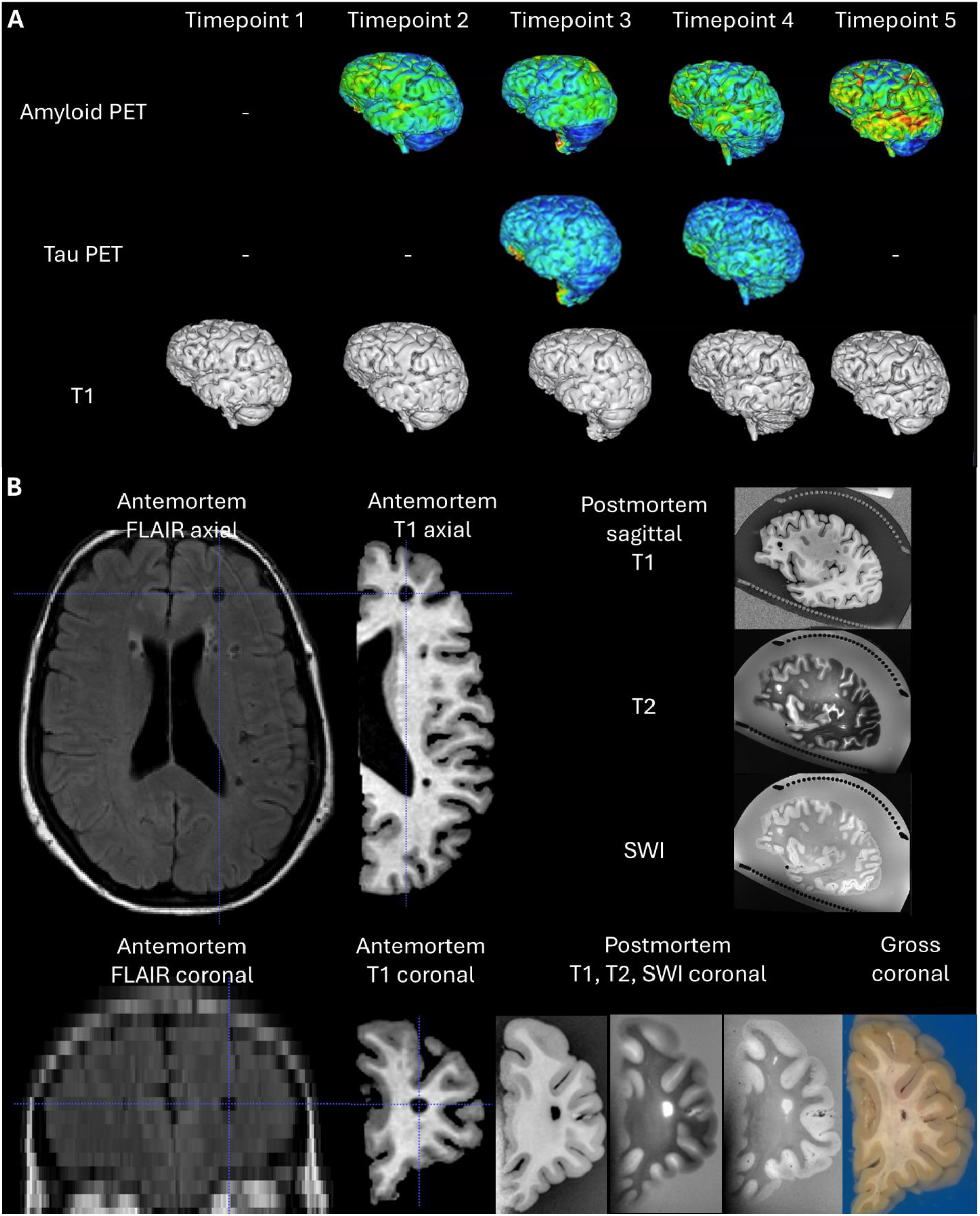
(A) Representative images of longitudinal amyloid PET, tau PET, and MRI. (B) Numerous cystic appearing lesions detected in structural MRI were hypointense on both T1 and T2 FLAIR, with subtle peripheral T2 signal abnormality involving many of the lesions. By aligning antemortem-postmortem-gross images, regions with high amyloid and tau burden can be analyzed.

Postmortem MRI of the left hemisphere revealed a hippocampal volume of 2577 mm^3^ and an amygdala volume of 768 mm^3^, both notably exceeding the group average of 2138 ± 611 mm^3^ and 401 ± 198 mm^3^, respectively. Based on the PET neuroimaging data, there may be a subtle increase in amyloid burden, particularly in the posterior cingulate, closer to the time of death, accompanied by minimal atrophy.

### Neuropathology

The fresh (whole) brain weight was 1083 g, exceeding the group average of 1043 ± 249 g. Grossly, mild atrophy was observed in the frontal cortex, parietal and temporal cortex, and hippocampus; however, this assessment may not be as precise as radiologic volumetric analysis.

Immunocytochemistry determined that this case was Thal phase 3 for amyloid beta deposition (A2) (Figure 2A-C), Braak stage 4 for neurofibrillary degeneration (B2) (Figure 2D-E), and a moderate CERAD score for neocortical neuritic plaques (C2) (Figure 2F), resulting in an intermediate AD (A2B2C2) neuropathologic change classification according to diagnostic criteria. Secondary neuropathology included Lewy body pathology and cerebrovascular pathology. Specifically, amygdala-predominant Lewy body pathology was identified (Figure 2G). Additionally, moderate cerebral amyloid angiopathy, notably affecting the gray matter (Figure 2H), was noted. Microcalcifications were observed in the basal ganglia, thalamus, and cerebellum (Figure 2I), along with mild arteriolosclerosis in the inferior parietal and midbrain regions (Figure 2J-K). Furthermore, microhemorrhage and macrophages in the medulla were observed (Figure 2L). Notably, no TDP-43 immunosignal was detected, and hippocampal sclerosis was absent. Overall, the main diagnosis for this individual with DS in her 60’s and a non-carrier status for ApoE4 was intermediate AD neuropathologic change, with secondary diagnoses of Lewy body pathology and cerebrovascular pathology.

**Figure 2.**
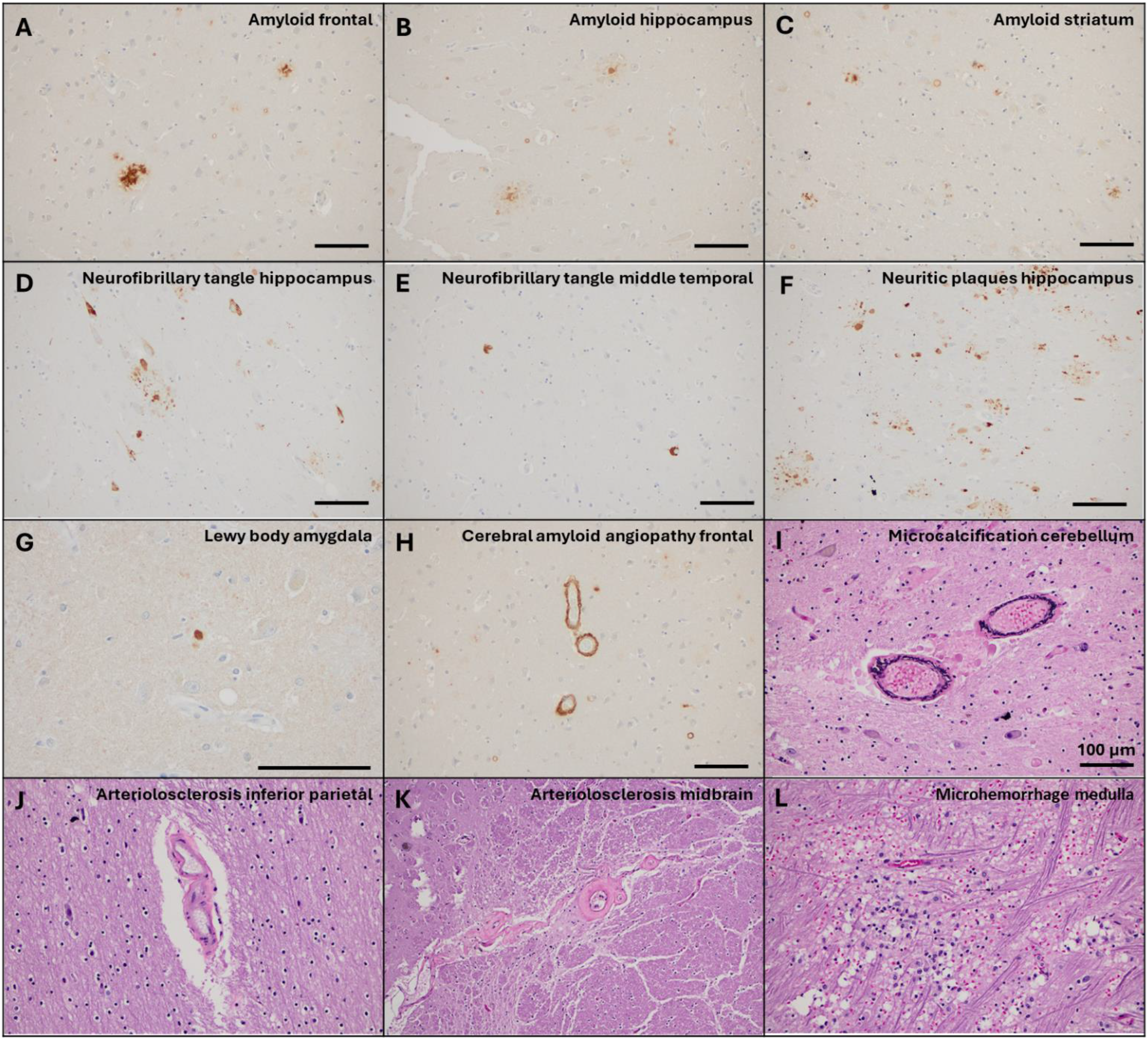
At autopsy, (A-C) beta-amyloid deposition is observed in the frontal cortex and the hippocampal CA1 as well as the striatum, leading to a diagnosis of Thal phase 3. Additionally, (D-E) neurofibrillary tangles are detected in the hippocampus and middle temporal gyrus, corresponding to Braak stage 4. (F) Nine neuritic plaques are detected in the hippocampus – a moderate CERAD score. (G) Lewy body pathology is evident in the amygdala. Cerebrovascular pathology includes (H) cerebral amyloid angiopathy in the frontal cortex, (I) microcalcification in the cerebellum, (J-K) arteriolosclerosis in the inferior parietal and midbrain, and (L) microhemorrhage in the medulla.

## Discussion

We describe a unique individual with DS, who had medical and cognitive test records over a 31-year period, and neuroimaging and fluid biomarker data during the last 10 years of her life. She maintained intact cognitive functioning despite showing intermediate AD neuropathology at autopsy. This case description contributes to the other reports of individuals with DS who did not develop dementia into their late 60s [52], 70s [53-57], and 80s [58].

This case contributes to the expanding body of literature [59] aimed at understanding the phenomenon of resistance to cognitive decline among individuals with AD neuropathology who remain non-demented. Various compensatory mechanisms have been proposed, including the concept of “cognitive reserve.” Research suggests that individuals with higher levels of education tend to exhibit less cognitive impairment compared to those with lower education levels, potentially due to more advantageous lifestyle choices [60-66]. In our case, the individual possessed notably high levels of education compared to peers with DS from her generation, suggesting a potential cognitive reserve effect.

Another proposed compensatory mechanism is “brain reserve.” Individuals with larger brain reserves, characterized by greater synaptic density and a larger number of healthy neurons, may require more pathology to exhibit clinical dementia symptoms [67, 68]. This delay in symptom manifestation may occur because substantial pathology accumulates before clinical symptoms become evident [69-72]. Our case exhibited a larger brain compared to typical individuals with DS in autopsy cases, suggesting a potential brain reserve effect.

A third potential compensatory mechanism is “genetic advantage.” Certain genetic mutations may confer resistance to dementia by inhibiting the formation of neurofibrillary tangles or enhancing the clearance of amyloid beta [73, 74]. This genetic advantage may be particularly relevant in cases of non-demented individuals demonstrating intermediate AD neuropathologic changes (NDAD), characterized by Braak stage 2-4 and moderate to frequent CERAD scores. Our case aligns with this profile, exhibiting features consistent with NDAD and intermediate AD neuropathologic changes.

In this case report, examination of longitudinal amyloid PET scans revealed an increase in cortical amyloid uptake compared to the cerebellum in several regions, including the superior frontal, rostral middle frontal, inferior parietal, and posterior cingulate areas. Tau PET ROIs exhibited a consistent level of tau accumulation, with SUVRs ranging from 1.0 to 1.4, indicating moderately elevated tau levels but remaining stable. However, it’s important to note that this assessment occurred three years before the autopsy. There is a possibility of a subsequent rapid accumulation of tau, potentially progressing towards Braak stage 4 for neurofibrillary degeneration (B2) as observed in neuropathology.

Despite being cognitively stable, neuropathological outcomes suggest AD pathology and amygdala-predominant Lewy body pathology. Lewy bodies in the amygdala in DS are not uncommon and depending on the cohort study prevalence ranges from 11% to 50% [75-78].

In elderly people [79], it is not uncommon to encounter cases where AD neuropathological changes are found in individuals clinically diagnosed as non-demented. For instance, in the London and Cardiff Brains for Dementia Research cohort, about 36% of cases showed such discrepancies, with 14% of clinically diagnosed controls exhibiting pathology at autopsy [80]. This case may be one of those instances where clinical and pathological diagnoses do not align.

In DS, there are 3 genetic causes including the most common full trisomy 21, mosaicism and partial trisomy [81-83]. A case report of partial trisomy 21 highlighted that the absence of APP overexpression was associated with a lack of AD development [57, 84]. Mosaicism for chromosome 21 at birth is linked to a phenotype characterized by less severe intellectual disability [85]. Since a blood karyotype was never completed, it is uncertain whether she had full trisomy 21 at birth. One possible explanation for her cognitive stability despite the presence of AD pathology is that she was full trisomy 21 at birth and acquired mosaicism throughout her life [27, 86]. Alternatively, she might have been mosaic at birth, with cognitive stability primarily influenced by other factors.

In contrast to other case reports detailing mosaic [58] and partial trisomy 21 [57], a recent study examined a 70-year-old man with complete trisomy 21 and ApoE3/3 [53], the longitudinal performance in mental status and episodic memory of this case was evaluated against that of one age-matched AD patient and one younger healthy participant. Despite exhibiting no symptoms of dementia, a gradual decline in visuospatial abilities was noted. Given his complete trisomy 21 and neutral ApoE genotype, along with a history of good health, the authors speculated that there might be genetic variations in gene expression associated with trisomy 21 or non-genetic factors such as learning and life experiences that ultimately contribute to successful aging and the avoidance of dementia.

The presence of ApoE4 leads to a high risk for AD in the general population while the presence of ApoE2 is thought to be protective [87-90]. In DS, multiple studies assessed the impact of ApoE genotype on the development of dementia. ApoE2 is linked to a decreased risk of dementia in individuals with DS [91-93]. In a large cohort study, the ApoE4 allele was associated with earlier clinical and biomarker changes of AD in DS. Thus, the presence of ApoE2/3 in this case study may also be part of the underlying reason for protection from dementia.

Factors like greater cognitive resilience may also have been protective in the person described here [94], as suggested by studies indicating that some individuals with DS maintain cognitive abilities through older ages [95, 96]. IQ or level of intellectual disability was not related to risk or age at onset of AD for adults with DS when severity of intellectual disability was included as a covariate in the analysis [97-99].

In DS, we have limited understanding of how to assess the impact of comorbidities in relation to biomarkers and other measures. Studies focusing on the late-onset AD population found that a higher Charlson comorbidity index, particularly BMI, is associated with elevated levels of plasma Aβ40 and tTau [100]. This association may explain why this participant exhibited higher plasma Aβ40 levels compared to the published studies (Table 2). Additionally, studies in the general population suggested that the increase in comorbidities is related to increasing age. For example, the prevalence of hearing impairment and thyroid disorders in DS dramatically increases after age 60 [101]. However, in contrast, a four-year prospective cohort study of neurotypical older adults with dementia found no association between Charlson comorbidity index and cognitive function [102].

A recent study investigating the correlation between inflammatory conditions and the age of AD onset in individuals with DS highlighted a potentially “protective” effect associated with a history of gout [103].The study, which examined 339 adults with DS, revealed a statistically significant delay in the onset of AD by approximately 2.5 years among those with a history of gout. This delay was theorized to potentially arise from the antioxidant properties of hyperuricemia. In the case at hand, there was no formal diagnosis of gout, nor any indication of medication typically prescribed for gout management. However, the participant did report arthritis in her knees and experienced pain in her big toe. While these symptoms could suggest gout, especially considering the typically high pain tolerance observed in individuals with DS, a definitive diagnosis requires confirmatory uric acid level testing. Although intriguing, this scenario underscores the importance of considering a history of gout or hyperuricemia in the clinical evaluation of individuals with DS. While such conditions may not always be apparent, they have the potential to impact the onset and progression of AD within this population.

Interestingly, there was a hint of diminished social functioning observed in the DLD sum of social scores in the last 3-4 years of her life . One plausible explanation could be the presence of Lewy bodies in the amygdala of this individual. However, it’s important to consider that the elevated sum of social scores over the two-year span likely stemmed from heightened agitation and interpersonal conflicts experienced with peers at her day program and her independent living services (ILS) supervisor. These challenges ultimately led to changes in her day program placements and ILS supervisor.

As more individuals with DS reach older ages, we anticipate identifying more that remain cognitively stable to allow for studies of resilience to AD neuropathology. Indeed, given the full penetrance of AD neuropathology over the age of 40 years in people with trisomy 21, this may be an exciting cohort to include in resilience studies.

## Data Availability

All data produced in the present work are contained in the manuscript

## Acknowledgements

The study team is grateful for the gift of brain donation by this ABC-DS participant. Funding to support this study was from NIH/NIA U19AG068054, P30AG066519, U01AG051412, RF1AG079519, P01AG025204, P01AG014449, and NIH/NICHD R01HD065160.

## List of abbreviations

Aβ40: amyloid beta 40
Aβ42: amyloid beta 42
AD: Alzheimer’s disease
ApoE: apolipoprotein E
APP: amyloid precursor protein
BMI: body mass index
CAA: cerebral amyloid angiopathy
CERAD: Consortium to Establish a Registry for Alzheimer’s disease
CS: cognitively stable
CSF: cerebrospinal fluid
DLD: Dementia Questionnaire for People with Learning Disabilities
DS: Down syndrome
DSMSE: Down Syndrome Mental Status Examination
FBP: florbetapir
FSIQ: full scale IQ
GFAP: glial fibrillary acidic protein
KBIT-2: Kaufman Brief Intelligence Test, 2nd Edition
MCI: mild cognitive impairment
MRI: magnetic resonance imaging
NfL: neurofilament light
PET: positron emission tomography pTau181: phosphorylated tau 181
RADD-2: Rapid Assessment for Developmental Disabilities, 2nd Edition
ROI: region of interest
SUVR: standardized uptake value ratio
tTau: total tau
WAIS-R: Wechsler Adult Intelligence Scale, Revised Edition

